# Reduced Norepinephrine Transporter Binding in Parkinson’s disease with dopa Responsive Freezing Gait

**DOI:** 10.1101/2022.03.14.22272365

**Authors:** J. Lucas McKay, Jonathan Nye, Felicia C. Goldstein, Barbara Sommerfeld, Yoland Smith, David Weinshenker, Stewart A. Factor

## Abstract

Freezing of gait (FOG) is a major cause of falling and leads to loss of independence in Parkinson’s disease (PD). The pathophysiology of FOG is poorly understood – although there is a hypothesized link with NE systems. PD-FOG can present in levodopa-responsive and unresponsive forms.

We examined NE transporter (NET) binding via brain positron emission tomography (PET) to evaluate changes in NET density associated with FOG using the high affinity selective NET antagonist radioligand [11C]MeNER (2S,3S)(2-[α-(2-methoxyphenoxy)benzyl]morpholine) in N=52 parkinsonian patients. We used a rigorous levodopa challenge paradigm to characterize patients as non-freezing PD (NO-FOG, N=16), levodopa responsive freezing (OFF-FOG, N=10), levodopa-unresponsive freezing (ONOFF-FOG, N=21), and primary progressive freezing of gait (PP-FOG, N=5).

Linear mixed models identified significant reductions in whole brain NET binding in the OFF-FOG group compared to the NO-FOG group (−16.8%, P=0.021). Additional contrasts tested post-hoc identified trends toward increased NET expression in ONOFF-FOG vs. OFF-FOG (≈10%; P=0.123). Linear mixed models with interaction terms identified significantly reduced NET binding in right thalamus in the OFF-FOG group (P=0.038). A linear regression analysis identified an association between reduced NET binding and more severe NFOG-Q score only in the OFF-FOG group (P=0.022).

This is the first study to examine brain noradrenergic innervation using NET-PET in PD patients with and without FOG. Based on the normal regional distribution of noradrenergic innervation and pathological studies in the thalamus of PD patients, the implications of our findings suggest that noradrenergic limbic pathways may play a key role in OFF-FOG in PD. This finding could have implications for clinical subtyping of FOG as well as development of therapies.

## Introduction

Freezing of gait (FOG), defined as “brief, episodic absence or marked reduction of forward progression of the feet despite intention to walk”, is a common, disabling feature of Parkinson’s disease (PD)^1^. It is a major reason for falling and injury and leads to loss of independence and poor quality of life^2,3^. FOG is considered a cardinal feature of PD with independent progression^4^. The pathophysiology is poorly understood. Several system related theories have been proposed but the potential contribution of norepinephrine (NE) has historically received a great deal of interest and has led to a drug approval in one country for this purpose. NE is a catecholamine that has widespread distribution in the brain and appears to play a key role in arousal, attention, stress responses, mood, and cognition^5-8^.

The primary NE nucleus in brain is the locus coeruleus (LC), which projects to subcortical and cortical regions, including motor, premotor and associative regions of the frontal cortex, thalamus, hypothalamus, brainstem, amygdala, hippocampus, and cerebellum^9-13^. The LC is one of the earliest sites of α-synuclein pathology in the brain and undergoes catastrophic degeneration later in PD^14-17^. LC terminal degeneration precedes cell body loss^18^ and is associated with decreased tissue NE levels in LC projection regions^13,19^. Studies in animal models and patients indicate that the loss of LC-NE may exacerbate motor symptoms of PD, contribute to dopamine neuron degeneration, and attenuate the therapeutic effects of levodopa^20-25^.

The hypothesis linking NE to FOG is primarily based on data showing that L-DOPS (droxidopa), a synthetic precursor to NE, reduces the symptom complex. L-DOPS treatment produced marked to moderate improvement of FOG in 25% of patients in a study from the 1980’s^26,27^, and similar improvement was reported in atypical parkinsonian cases^28^. However, these findings are controversial and have not been replicated^4^. This work led to approval of droxidopa in Japan for FOG two decades ago. L-DOPS has also been examined and recently approved in the U.S. for orthostatic hypotension^29^. Other noradrenergic drugs including atomoxetine and stimulants appear to be effective in some patients with FOG^30-32^. Further, an association between FOG and NE deficit was shown in a study utilizing 6-[^18^F]fluoro-l-m-tyrosine (FMT) PET (a marker for NE metabolism). This study showed that decreased FMT uptake in LC was strongly correlated with increased self-reported FOG severity in PD patients^33^. This observation must be confirmed through more comprehensive studies in larger cohorts of patients. Finally, the relationship between FOG and anxiety is also consistent with the link between FOG and NE^34,35^. FOG can be elicited in situations of elevated stress^36^, which increases LC activity, NE release and arousal in animal models^6,37-39^. This suggests the hypothesis that a dysregulated LC-NE system may have aberrant responses to stress in PD that could, in turn, trigger FOG episodes.

In this study we examined NE transporter (NET) binding via positron emission tomography (PET) to evaluate changes in NET density associated with FOG. NET is a member of the Na+/Cl-dependent neurotransmitter transporter gene family located on the nerve terminals as well as the somatodendritic field of NE neurons and serves as a proxy for LC terminal integrity. We used the radioligand [11C]MeNER (2S,3S) (2-[α-(2-methoxyphenoxy) benzyl]morpholine), which has high affinity and is a selective antagonist for NET ^40,41^, to compare NET availability across PD patients with and without FOG, as well as in a small group of patients with non-PD FOG (referred to as primary progressive Freezing of Gait)^42^. We also separated PD-FOG patients into those responsive and non-responsive to levodopa^43^.

## Materials and Methods

### Study population

Study patients were recruited from the Emory Movement Disorders Clinic and provided written informed consent according to procedures approved by the Institutional Review Board of Emory University. Inclusion criteria for PD were: Age≥18 years; PD diagnosis defined by the United Kingdom Brain Bank criteria^44^; Hoehn & Yahr stage IV or higher in the OFF state; levodopa responsive; capable of signing a consent document and willing to participate in all aspects of the study. Inclusion criteria specific to participants with FOG were FOG noted in medical history and confirmed visually by examiner. Patients with primary progressive freezing gait (PP-FOG) diagnosed based on previously reported criteria^45^ were included; while patients previously treated with dopamine receptor blocking medications; or with neurological or orthopedic disorders interfering with gait including vascular parkinsonism; dementia or other medical problems precluding completion of study protocol were excluded.

### Levodopa challenge paradigm

The details of the levodopa paradigm challenge have been published elsewhere^43^. In brief, patients came to clinic in the practically defined “OFF” state >12 hours after last intake of antiparkinsonian medications^46^. They were assessed for motor symptoms using the Movement Disorder Society-Unified Parkinson’s Disease Rating Scale part III motor examination (MDS-UPDRS-III). After the assessment, patients were administered a levodopa equivalent dose (LED) greater than their typical morning dose. The assessment was repeated after patients reached their full “ON” state.

### FOG group assignment

We classified each PD patient into one of three study groups based on history of the presence of FOG along with scores on the MDS UPDRS-III item 11 “Freezing of Gait” exam in each of the “OFF” and “ON” medication states. Participants who had no history of FOG and received a score of zero on item 11 in both medication states were classified as “no freezing,” or “NO-FOG.” Participants who received a nonzero score on item 11 in the “OFF” medication state but a zero score in the “ON” medication state were classified as “OFF-FOG.” Participants who received a nonzero score on item 11 in both medication states were classified as “ONOFF-FOG.” Patients in the primary progressive Freezing of Gait group were classified as “PP-FOG.”

### [^11^C]MeNER-PET Imaging

Patients were assessed with NET-PET imaging on a separate testing day. The typical interval between levodopa challenge testing and NET-PET imaging was 2.2 ± 2.8 months. Urine pregnancy tests were obtained within 24 hours prior to injection of [^11^C]MeNER for female subjects, unless no menses during the past consecutive 12 months. Any noradrenergic medications were held for a duration based on the half-life. A telephone contact seven days post imaging session was made to assess for any adverse events. Patients maintained their typical medication schedule for NET-PET imaging.

#### Scanning Procedure

[^11^C]MeNER was synthesized by methylation of (S,S)-2-(α-(2-Methoxyphenoxy)benzyl)morpholine (MeNER), an analogue of the selective norepinephrine reuptake inhibitor (S,S)-reboxetine, as previously described^47^. The administered activity ranged from 570 to 832 MBq delivered intravenously with an average specific activity of 32 ± 12 GBq/µmol, resulting in an injected mass ranging between 3.4 and 17 µg.

#### Imaging Procedures

PET data was collected on a High-Resolution Research Tomography (CTI, Inc. Knoxville, TN) at the Emory Center for Systems Imaging. The PET system has a 2.5mm spatial resolution and 10-fold higher sensitivity than current clinical PET cameras^48^. Subjects had a venous catheter placed in their forearm and were administered [^11^C]MENeR in a quiet uptake room for 60 minutes. Subjects were then placed supine on the imaging table with their head lightly restrained using Velcro straps, and 30 min of PET data was collected. PET data were divided into six frames, each five minutes duration and reconstructed with an ordinary Poisson ordered-subset expectation maximization (OP-OSEM) algorithm (6 iterations, 16 subsets) including scatter and attenuation correction.

Structural MRI images were acquired on a 3T Siemens Prisma scanner with a 32-channel receiver array head coil. A standard whole-brain 3D T1-weighted MPRAGE sequence^49^ recommended for brain structure morphometric analysis^50^ (FOV = 256 mm; TR/TI/TE/FA = 2530 ms/1100 ms/3 ms/8°; 1 mm X 1 mm X 1 mm resolution; time of acquisition ∼ 9 minutes) was acquired to provide anatomic detail. Foam padding was utilized to minimize head motion. These data were be used for normalization of PET data to the standard Montreal Neurological Institute analysis space.

#### Image Analysis

PET data was corrected for inter-subject motion and co-registered to their structural MRI scan by optimization of the mutual information metric. MRI structural data were then spatially normalized to the Montreal Neurological Institute (MNI T2 152) template with SPM8’s segment option and default parameters. The inverse non-linear transform was used to map regions of interest from the Anatomical Automatic Labeling template^51^ including the frontal lobe, left and right thalamus, temporal lobe, LC and cerebellum. Regional standardized uptake value ratios (SUVR) were extracted from the reconstructed PET data normalized to the cerebellum^33^. We have shown that this approach is highly correlated with NET binding density in structurally similar analogue based on reboxitine^52^. Imaging technologists and those assessing results were blinded to the their group assignment.

### Statistical Analysis

All summary statistics are presented as mean (standard deviation) or frequency (percentage). Clinical and demographic variables were compared across study groups using univariate tests (Chi-squared, ANOVA). Variation in NET binding across study groups was assessed with a linear mixed model with fixed effects for study group (NO-FOG, OFF-FOG, ONOFF-FOG, and PP-FOG, with NO-FOG as the reference group) and brain region (Frontal cortices, left thalamus, right thalamus, locus ceruleus, temporal lobes, with frontal cortices as the reference group) and a random effect for patient. Subsequent models evaluated sensitivity of main results to inclusion of covariates identified as marginally significant in univariate tests (sex, duration). Region-specific group effects were assessed with an additional linear mixed model with region by group interaction terms. Linear mixed models were implemented in *LmerTest::lme4* in R software and fitted using restricted maximum likelihood. Linear regression models were applied to evaluate associations between NET expression and self-reported FOG severity (NFOG-Q) score in regions identified as varying with FOG group (*stats::lm* in R software). All statistical analyses were performed in *R* v4.1.1 at alpha = 0.05.

### Data availability

The datasets generated during and/or analyzed during the current study are available from the corresponding author on reasonable request.

## Results

### Clinical and demographic characteristics

A total of N=60 patients were enrolled and assessed with levodopa challenge. Of these, PET data was subsequently obtained for 52: Sixteen were classified as NO-FOG, 10 as OFF-FOG, and 21 as ONOFF-FOG. The remaining five patients were PP-FOG. Clinical and demographic characteristics are presented in Table 1. No statistically significant differences in frequency of missing data were observed across study groups (**Supplemental Information**). No statistically significant differences in clinical or demographic variables were observed between groups; with the exception of N-FOGQ score and MDS-UPDRS-III FOG item scores (expected from the design) and marginally significant effects of sex (P=0.05) and disease duration (P=0.08). Higher levels of anxiety were reported among those with freezing (average scores of 10.8±8.2 vs. 8.4±6.1 on the Beck Anxiety Inventory); however, this contrast was not statistically significant (P=0.69, ANOVA).

**Table 1.** Demographic and clinical characteristics of the study sample.

| Variable | NO-FOG<br>(N=16) | OFF-FOG<br>(N=10) | ONOFF-FOG<br>(N=21) | PP-FOG<br>(N=5) | Total (N=52) | P Value |
| --- | --- | --- | --- | --- | --- | --- |
| Age |  |  |  |  |  | 0.88 |
| Mean (SD) | 67.3 (11.8) | 68.5 (5.2) | 68.6 (6.6) | 65.6 (5.9) | 67.9 (8.2) |  |
| Sex |  |  |  |  |  | 0.05 |
| Female, N (%) | 5 (31%) | 1 (10%) | 2 (10%) | 3 (60%) | 11 (21%) |  |
| Male, N (%) | 11 (69%) | 9 (90%) | 19 (90%) | 2 (40%) | 41 (79%) |  |
| Duration |  |  |  |  |  | 0.08 |
| Mean (SD) | 6.0 (3.7) | 10.2 (5.4) | 10.2 (6.9) | 6.0 (3.3) | 8.5 (5.8) |  |
| MDS-UPDRS-III (OFF) |  |  |  |  |  | 0.45 |
| Mean (SD) | 31.1 (13.6) | 30.6 (10.7) | 34.3 (10.8) | 39.4 (7.8) | 33.1 (11.5) |  |
| MDS-UPDRS-III (ON) |  |  |  |  |  | 0.08 |
| Mean (SD) | 18.4 (14.9) | 15.9 (8.1) | 20.5 (9.1) | 31.6 (9.0) | 20.0 (11.6) |  |
| FOG Score (OFF) Item 11 |  |  |  |  |  | < 0.01 |
| 0, N (%) | 16 (100%) | 4 (40%) | 3 (14%) | 2 (40%) | 25 (48%) |  |
| 1-3 | 0 (0%) | 6 (60%) | 15 (71%) | 2 (40%) | 23 (44%) |  |
| 4 | 0 (0%) | 0 (0%) | 3 (14%) | 1 (20%) | 4 (8%) |  |
| FOG Score (ON) Item 11 |  |  |  |  |  | 0.029 |
| 0, N (%) | 16 (100%) | 10 (100%) | 16 (76%) | 2 (40%) | 44 (85%) |  |
| 1-3 | 0 (0%) | 0 (0%) | 4 (19%) | 2 (40%) | 6 (12%) |  |
| 4 | 0 (0%) | 0 (0%) | 1 (5%) | 1 (20%) | 2 (4%) |  |
| NFOG-Q |  |  |  |  |  | < 0.01 |
| Mean (SD) | 0.0 (0.0) | 16.7 (6.2) | 21.2 (3.7) | 17.8 (7.5) | 13.5 (10.1) |  |
| MoCA |  |  |  |  |  | 0.30 |
| Mean (SD) | 26.1 (3.9) | 24.3 (4.0) | 23.3 (5.0) | 25.2 (4.3) | 24.6 (4.5) |  |
| BAI |  |  |  |  |  | 0.69 |
| Mean (SD) | 8.4 (6.1) | 10.5 (7.6) | 10.4 (8.2)<sup>a</sup> | 12.8 (10.6) | 10.0 (7.6)<sup>b</sup> |  |

MDS-UPDRS-III, Unified Parkinson's Disease Rating Scale, Movement Disorders Society revision; Item 11 from part III of MDS-UPDRS; MoCA, Montreal Cognitive Assessment; BAI, Beck Anxiety Inventory. <sup>a</sup>N=20; <sup>b</sup>N=51.

### NET binding

In general, a similar pattern of variation in NET binding across study groups was observed across all brain regions of interest. The lowest levels of NET SUVR values were observed in the OFF-FOG group, followed by PP-FOG, ONOFF-FOG, and NO-FOG, respectively. (Region-specific values are summarized in **Supplemental Information; Table S1**). Linear mixed models identified significant reductions in whole brain NET binding in the OFF-FOG group compared to the NO-FOG group (change in SUVR -16.8%, P=0.021; Figure 1). Additional contrasts tested post-hoc identified trends toward increased NET binding in ONOFF-FOG vs. OFF-FOG (≈10%; P=0.123). No trends were observed for the PP-FOG group. Models controlling for sex and disease duration produced very similar results (−16.1% and -14.4%, respectively), although the contrast between OFF-FOG and NO-FOG was only marginally significant (P=0.054) when controlling for duration. Linear mixed models with interaction terms between study group and brain region identified a significant reduction in NET binding in right thalamus in the OFF-FOG group (P=0.038). A similar reduction of smaller magnitude was observed in left thalamus (P=0.157).

**Figure 1.**
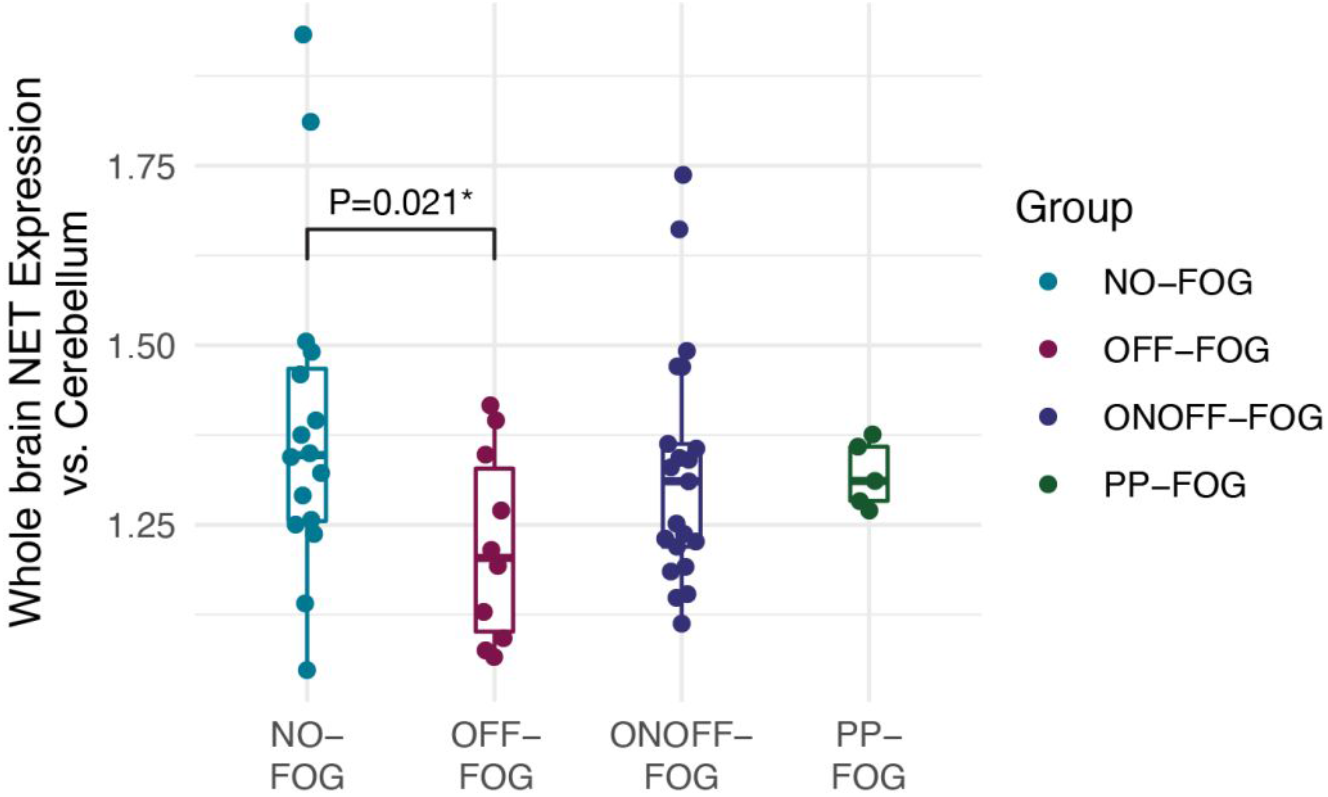
Variation in NET binding with FOG classification. A linear mixed model identified statistically significant reduction in NET binding across all brain regions in the OFF-FOG group vs. the NO-FOG group.

### Right thalamus NET expression and NFOG-Q score

Because of the strong interaction observed between NET binding and OFF-FOG group in the right thalamus, we tested whether NET abundance in this area was associated with self-reported FOG severity as measured on NFOG-Q. The PP-FOG group was excluded from this analysis due to the small number of cases. A linear regression of NFOG-Q score onto right thalamus NET availability identified a statistically significant negative relationship, with reduced NET associated with more severe NFOG-Q score in the OFF-FOG group (P=0.022), but not in the other groups (Figure 2). The overall variation in NFOG-Q score explained by NET binding with interaction terms for group was *R*^*2*^_*adj*_ = 0.88. Similar relationships were observed in a model with the square root of NFOG-Q score as the dependent variable to stabilize variance (**Supplemental Information**).

**Figure 2.**
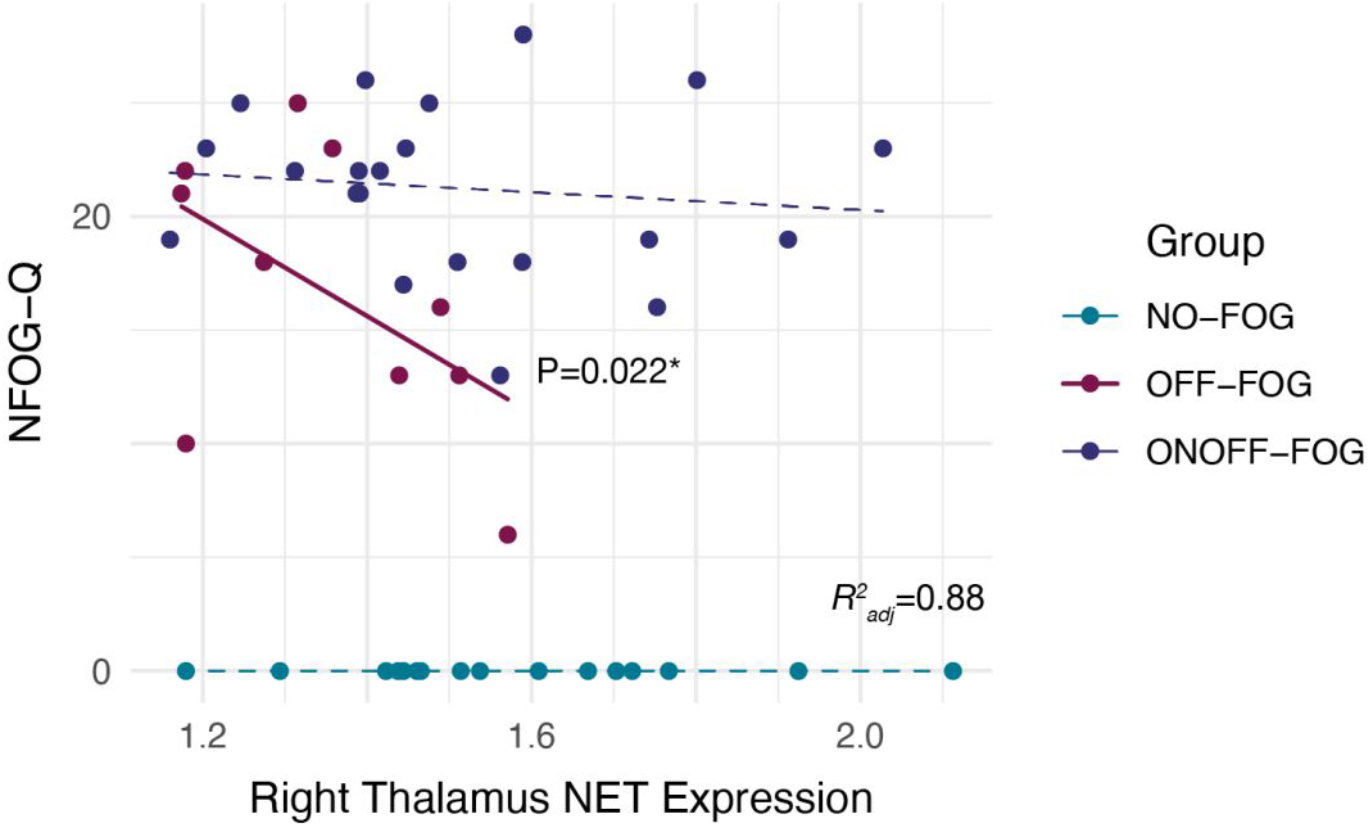
Association between NET binding in right thalamus and NFOG-Q score. Linear regression identified a statistically significant association between NET binding and NFOG-Q among OFF-FOG (solid line) but not among the other groups (dashed lines).

### Additional analyses

Similar results were obtained with additional linear mixed models performed post-hoc that: 1) considered only those cases with imaging within 3 months of levodopa challenge (NET binding in OFF-FOG vs. NO-FOG, -16.0%, P=0.043), 2) that excluded the PP-FOG group (−16.8%, P=0.027), and 3) that excluded the LC (−17.2%, P=0.020).

## Discussion

There is mounting evidence supporting a role for extranigral noradrenergic (NA) systems in the development of FOG in PD. This is a first look at the use of NET-PET imaging using the ligand [^11^C]MeNER as a tool to examine the role of NA denervation in PD-FOG. We examined whole brain NET binding via PET to evaluate changes in NET density associated with FOG; and particularly, in relation to FOG responsiveness to levodopa. We scanned 47 PD patients (16 NO-FOG, 10 OFF-FOG, 21 ONOFF-FOG) and 5 patients with primary progressive freezing gait (PP-FOG). We found significantly decreased whole-brain NET binding in the OFF-FOG group compared to NO-FOG (P=0.021). Importantly, we showed that this deficit was specific to OFF-FOG; patients with ONOFF-FOG or PP-FOG showed NET binding comparable to those without FOG. Additional region-specific comparisons revealed decreased binding in the right thalamus (P=0.038) with a trend for the left thalamus (P=0.16), in the PD OFF-FOG group. These findings support the hypothesis that a loss of NA innervation, perhaps particularly to the thalamus, may contribute to levodopa responsive FOG (OFF-FOG) in PD patients. The results also add to previous findings suggesting a connection between NA dysfunction and FOG. Further, this decreased NA innervation may relate to the recently reported increase in CSF Aβ42 amyloid in PD-FOG patients^53^ as previously suggested by studies in non-human primates and several transgenic rodent models where the reduction of NE contributed to increased Aβ pathology^54-56^. Interestingly, the increased amyloid aggregation in these models was reversed by using droxidopa^57^.

### Structural and functional changes in the thalamus in FOG

This work adds to a growing body of knowledge demonstrating structural and functional changes in the thalamus – and in the connections of the thalamus to other brain regions – in PD patients with FOG. In a prospective MRI neuroimaging study, D’Cruz et al^58^ showed local inflations in bilateral thalami in patients who presented with or would later develop FOG. These inflations involved medial thalamic sub-nuclei volumes which are highly innervated by NA pathways. Resting-state analyses at baseline further revealed that patients who would go on to develop FOG had thalamo-cortical coupling with limbic and cognitive regions initially stronger than that among patients who would not go on to develop FOG. This initial increased coupling exhibited a marked reduction over the two years. The authors hypothesized that many of these changes may have been compensatory in nature, with increased reliance on non-motor thalamic networks, particularly limbic connections, in PD patients with FOG; after initial success, decompensation occurred, and FOG would emerge over the course of disease progression.

Other studies have shown several anatomical changes in connections between the thalamus and other brain regions in FOG, including some known to be crucial for balance and gait. These include diminished structural connectivity between the thalamus and mesencephalic locomotor region and reduced thalamic fiber tracts in the right hemisphere in FOG^59^ and more pronounced white matter abnormalities in multiple tracts connecting to the thalamus in FOG^60^. It has also been shown that FOG episodes are associated with decreased anterior thalamic Blood-Oxygen-Level-Dependent (BOLD) signal, potentially associated with paroxysmal reductions in mesencephalic locomotor region activity^61,62^.

Our results imply that there is decreased NA innervation of the thalamus in patients with levodopa responsive FOG, which strongly suggests that interaction between NA sensory signaling and dopaminergic movement regulation is particularly important for OFF-FOG phenotype. It is possible that in the presence of dopamine replacement, some patients are able to successfully leverage non-motor regions of basal-ganglia-thalamocortical loops to compensate for impaired NA motor – specifically gait and balance – pathways. It is conceivable that this could lead to compensatory “inflation” of these areas as described by D’Cruz and colleagues, and that additional damage to other tracts impinging on the thalamus would impact this compensation. However, because the majority of these studies did not classify patients by FOG levodopa responsive state, it is difficult to draw firm conclusions. What appears clear is that there is a class of patients with greater NA deficits in the thalamus who do not experience FOG in the presence of adequate dopamine.

### Noradrenergic innervation of the thalamus

Noradrenergic innervation of the thalamus has been examined in monkey and human in some detail. In a study of two nonhuman primates, NA axon maps were generated with dopamine *β-*hydroxylase (DβH) and NET immunohistochemistry, and in vitro quantitative autoradiography for alpha-1 and alpha-2 adrenergic receptor densities^63^. The distribution patterns of DβH, NET, and adrenergic receptors were very similar with some modest variation noted. The most densely innervated thalamic regions were the midline, caudal intralaminar (paracentral and parafascicular), and medial mediodorsal nuclei. NA axons were present at moderate levels within the anterior nuclei, while the ventral motor thalamic nuclei exhibited moderate to low NA innervation. The sensory relay nuclei were generally less innervated. The visual relay nucleus, dorsal geniculate nucleus displayed the lowest NA innervation. These findings were concordant with postmortem human results^13,64,65^.

Although NET-PET does not provide the spatial resolution to distinguish the distribution of NET binding within the various thalamic nuclei, the impact of decreased thalamic NET binding can be speculated based on these findings and considering the systems thought to be involved in freezing, including motor, visual, cognitive, and limbic circuits^66^. It is noteworthy that the highly NA-innervated caudal intralaminar regions include predominantly thalamic neurons that project heavily to associative and limbic striatum^67,68^. The dense NA axonal innervation of these thalamic nuclei may allow for an indirect NA modulation of basal ganglia activity across motor and non-motor functional domains^63^. Furthermore, these densely innervated regions are known to receive extensive cholinergic innervation from the pedunculopontine tegmental area^64^, a cholinergic brain network that is also severely disrupted in PD^69^.

### T<u>halamic Pathology in PD</u>

Thalamic pathology has been studied in PD using neurochemical and anatomic methods. Unfortunately, none of these studies examined PD-FOG separately. Nevertheless, they provide some important relevant information. Pifl et al^13^ reported widespread loss of thalamic NE including motor (>85%), limbic (>/= 78%), and intralaminar/midline nuclei in PD compared to controls. The widespread nature of the loss of thalamic NE in PD is likely the result of the well-established degeneration of the LC^17^. Rub et al ^70^ showed that areas of increased Lewy inclusion pathology in the thalamus roughly corresponded to areas of dense NA innervation. These findings indicate that the burden of the PD-related pathology is seen in nuclei representing essential parts of the motor and limbic circuit^70^.

### Limbic circuits and FOG

These results are consistent with the hypothesis that abnormal limbic-striatum connectivity may interfere with processing specific anxiety provoking situations that lead to FOG – particularly OFF-FOG. As shown, the thalamic NA system is closely linked to limbic circuits and, in turn, we have shown in this study a link between NA changes and FOG. Other studies have shown a connection between FOG and limbic circuit changes. The limbic circuit plays a critical role in generating adequate emotional and behavioral responses, regulation of motivational processes and social behavior, and emotional processing, particularly of stimuli that induce anxiety and depression, including in PD^71-75^, all of which are essential for learning and memory. It has been suggested that this circuit is connected to the development of FOG based on several reports of worsening anxiety and depression in PD-FOG patients compared to PD-NOFOG^34,35^. Further, greater situational anxiety is also associated with worse FOG severity^35^. Imaging studies ^66,76^ have found that connectivity between the amygdala and striatum was significantly increased in freezers compared to non-freezers. It is suggested that the increased connectivity between the limbic circuit and striatum represents over-integration between these normally segregated networks which may lead to transient overload of motor network processing, that is the already dopamine depleted, leading to transient increased inhibitory input of the basal ganglion leading to motor breakdown and freezing^77,78^. In addition, anti-coupling between the cortical and subcortical limbic network and within the subcortical network was marked during freezing^66^. This might suggest there is an abnormality in processing specific anxiety provoking situations that lead to FOG, particularly OFF-FOG, and that this relates to abnormal limbic-striatum connectivity. Considering that anxiety and panic frequently occur as manifestations of off episodes^79^ it makes sense that the limbic circuitry would relate to the OFF-FOG group specifically.

### ONOFF vs OFF FOG in PD: Different pathophysiology?

The results of this study provide additional evidence supporting the hypothesis that OFF-FOG and ONOFF-FOG result from distinct pathophysiology. We have shown that OFF-FOG relates to hallucinatory phenomena while ONOFF-FOG is associated with distinct cognitive features – with the implication that these two types are associated with distinct pathophysiology in regions outside the basal ganglia^80^. We have also shown that axial and lower limb parkinsonian signs are also less severe and more levodopa responsive in OFF-FOG than ONOFF-FOG – with the implication that these types are associated with distinct pathophysiology within the basal ganglia^43^. The present results also support the notion that OFF-FOG and ONOFF-FOG are likely governed by separate pathophysiological mechanisms.

We have now shown a potential neurochemical marker separating FOG levodopa responsive subtypes, whereby only the OFF-FOG group demonstrates significantly lower NA innervation. Although some investigators have suggested that FOG progresses through a continuum from OFF-FOG to ONOFF-FOG – indicating that they are not separate entities^81^ – others support the idea that ONOFF-FOG is an independent form that can develop without prior OFF-FOG^82,83^. In fact, we previously showed through detailed examination of medical records that ONOFF-FOG can appear without first transitioning through OFF-FOG^43^. Our results provide neurochemical support that OFF-FOG is distinct from ONOFF-FOG, as it is unlikely that over time NA innervation in the thalamus would increase in OFF-FOG patients with the development of ONOFF-FOG.

### The potential of noradrenergic drugs for PD OFF-FOG

These results might suggest that NA agents would be of greater use in the subgroup of PD patients with “OFF-FOG.” While these patients are responsive to levodopa, it is often at the cost of greater dyskinesia^43^. While NA antagonist drugs can alleviate levodopa-induced dyskinesia in some animal models and patients^84-88^, the impact on causing or worsening dyskinesia is likely limited. This would make use of NA drugs reasonable alternatives. Several NA medications have been examined in a limited fashion for PD-FOG. Droxidopa was shown in a large double-blind, placebo control trial (n=218) in Japan in the 1980’s to moderately to markedly improve freezing in 25% of PD patients with FOG^26^. This led to approval of droxidopa for FOG in that country. The effectiveness of this drug has not been replicated^4^. There have also been two small pilot studies reported by Jankovic et al (n=5) and Revuelta et al (n=10) examining atomoxetine for the treatment of FOG, both showed modest benefits^30,32^. Finally, a multi-center, parallel-group, double-blind, placebo-controlled, randomized trial of methylphenidate (1 mg/kg per day for 90 days) for FOG in PD patients previously treated with subthalamic nucleus deep brain stimulation demonstrated that methylphenidate improved gait hypokinesia and freezing^31^.

Based on these results, we speculate that trials of NA agents for FOG would likely benefit from including only those patients with OFF-FOG (those with greatest decrease in NET PET signal), as it stands to reason that only these patients will likely benefit. Previous trials may have underestimated efficacy of agents if ONOFF-FOG cases were included. We consider this to be relatively likely, as patients with more severe FOG often have unresponsive FOG. Therefore, those who might have been the most likely to be recruited for trials might have been those least likely to benefit.

### Non-PD FOG

In this study we included a separate group of five subjects with primary progressive freezing gait disorder (PP-FOG). Overall, the results suggest that degeneration of NA system may not significantly contribute to PP-FOG. PP-FOG is a syndrome characterized by early freezing, bradykinesia, no response to levodopa and a stereotyped course of progression^45^. The syndrome can be the result of several disorders, most commonly progressive supranuclear palsy^42,89^. Relatively little is known about this syndrome regarding pathology, pathophysiology, and neurochemistry. It is known that not all PP-FOG patients have nigrostriatal dopaminergic degeneration^89^, and it has been suggested that PP-FOG may result from extranigral neuronal degeneration. Based on our results, we suggest that disruption of the NA system may not contribute significantly to this type of FOG, because NET-PET binding was similar to the ONOFF-FOG and NO-FOG groups but differed from the OFF-FOG group. However, because of the small sample size, additional subjects will be needed to confirm this result, taking into consideration that multiple pathophysiological bases exist for ONOFF-FOG.

### Limitations

There are some limitations to this study. The sample size was moderate, but due to the well-recognized demographic and clinical imbalances in PD patients with and without FOG, it remains difficult to completely control for potential confounds related to increased disease progression and age usually observed in FOG patients. We are encouraged by the fact that the main deficit seen in OFF-FOG was not seen in more severe ONOFF-FOG, which makes confounding by overall disease severity less likely. Because of the nature of observational studies, the potential for confounds remain, and these results should be replicated in a larger sample and preferably in a prospective context. In addition, spatial resolution of PET has reduced our ability to image certain regions such as specific nuclei of the thalamus and the LC.

## Conclusion

This is the first study to examine brain NA innervation using NET-PET in PD patients with and without FOG. We found significantly decreased whole-brain NET binding in the OFF-FOG group compared to NO-FOG. Additional region-specific comparisons revealed decreased binding in the thalami, right greater than left and more notable for the PD OFF-FOG group than the ONOFF-FOG and NO-FOG groups. Based on the normal regional distribution of NA innervation and pathological studies in the thalamus of PD patients, our findings suggest that limbic pathways may play a key role in OFF-FOG in PD. This finding could have implications for clinical subtyping of FOG as well as development of therapies.

## Data Availability

All data produced in the present study are available upon reasonable request to the authors

## Abbreviations

DβH: dopamine *β-*hydroxylase ()
FMT: 6-[^18^F]fluoro-l-m-tyrosine
FOG: Freezing of gait
LED: levodopa equivalent dose
LC: Locus Ceruleus
MNI: Montreal Neurological Institute
MDS-UPDRS-III: Movement Disorder Society-Unified Parkinson’s Disease Rating Scale part III motor examination
NA: noradrenergic
NE: norepinephrine
NET: Norepinephrine Transporter
N-FOGQ: New FOG questionnaire
as NO-FOG: no freezing
OFF-FOG: those with FOG in OFF state only
ONOFF-FOG: those with FOG in the ON and OFF state
PD: Parkinson’s disease
PP-FOG: primary progressive freezing gait
SUVR: standardized uptake value ratios

## Supplementary material

‘Supplementary material is available at Brain online’.

## Supplemental Information

### Incomplete imaging data

Of N=60 participants originally enrolled in the study, NET-PET imaging was not completed for N=8 (N=1, NO-FOG; N=1, OFF-FOG; N=5, ONOFF-FOG; N=1, PP-FOG). Only partial documentation for missing data was available. Reasons for missing data were: patient moved away, N=1 NO-FOG; inability to place head in scanner, N=1 OFF-FOG, no documentation available, N = 5 ONOFF-FOG, N=1 PP-FOG. No statistically significant difference in frequency of missing imaging data was observed across study groups (P=0.61, chi-squared test).

### Numerical values of NET expression

Regional standardized uptake value ratios (SUVR) extracted from the reconstructed PET data normalized to the cerebellum are shown in Table S1. Numerical values of 1.0 correspond to average expression equal to that observed in the cerebellum; numerical values ≥ 1.0 correspond to average expression greater than that observed in the cerebellum, which is considered to have negligible NET expression.

**Table S1.** Numerical NET SUVR values by study group.

| Region | NO-FOG<br>(N=16) | OFF-FOG<br>(N=10) | ONOFF-FOG<br>(N=21) | PP-FOG<br>(N=5) | Total<br>(N=52) |
| --- | --- | --- | --- | --- | --- |
| Locus Ceruleus | 1.28 (0.27) | 1.13 (0.25) | 1.23 (0.18) | 1.23 (0.09) | 1.23 (0.22) |
| Frontal Cortices | 1.20 (0.22) | 1.09 (0.12) | 1.15 (0.15) | 1.12 (0.03) | 1.15 (0.17) |
| Temporal Lobe | 1.31 (0.22) | 1.15 (0.09) | 1.24 (0.12) | 1.25 (0.03) | 1.25 (0.16) |
| Thalamus (Left) | 1.56 (0.27) | 1.37 (0.14) | 1.49 (0.24) | 1.49 (0.12) | 1.49 (0.23) |
| Thalamus (Right) | 1.58 (0.23) | 1.35 (0.15) | 1.51 (0.23) | 1.51 (0.07) | 1.50 (0.22) |
| Grand Mean | 1.39 (0.22) | 1.22 (0.13) | 1.32 (0.16) | 1.32 (0.05) | 1.32 (0.18) |

### Linear regression of NFOG-Q score onto right thalamus NET expression with square root transform

Because of the potential that non-normal distribution of residuals might invalidate the assumptions of linear regression, we repeated the regression analysis using square-root transformed NFOG-Q scores as the outcome variable. The results were very similar to those of the linear regression model presented in the main body of the manuscript, with a significant variation in NFOG-Q score with right thalamus NET expression in the OFF-FOG group (P=0.012) but not in the other groups. The overall variation in square root transformed NFOG-Q score explained by the model was *R*^*2*^_*adj*_ = 0.96.

### Association between MDS-UPDRS-III FOG score and right thalamus NET expression

We also examined whether right thalamus NET expression was associated with increased FOG severity as measured on the MDS-UPDRS-III. We dichotomized MDS-UPDRS-III FOG score in each of the ON and OFF states as 0-1 or ≥2 and dichotomized right thalamus NET expression about its median. Logistic regression models with terms for NET expression category and Group failed to identify statistically significant relationships in either the OFF or ON state.

